# The prevalence of scabies in Monrovia, Liberia: a population-based survey

**DOI:** 10.1101/2020.06.25.20139808

**Authors:** Shelui Collinson, Joseph Timothy, Samuel K Zayay, Karsor K Kollie, Eglantine Lebas, Katherine E Halliday, Rachel Pullan, Mosoka Fallah, Stephen L Walker, Michael Marks

## Abstract

**Background:** Scabies is known to be a public health problem in many settings but the majority of recent data is from rural settings in the Pacific. There is a need for high quality data from sub-Saharan Africa and peri-Urban settings to inform scale up of scabies control efforts. There have been anecdotal reports of scabies being a public health problem in Liberia but robust data are lacking.

**Methods:** We conducted a cross-sectional cluster-randomised prevalence survey for scabies in a peri-urban community in Monrovia, Liberia in February-March 2020. Participants underwent a standardised examination conducted by trained local health care workers. Health related quality of life (HRQoL) was assessed using age-appropriate dermatology life quality indices (DLQIs). Prevalence estimates were calculated accounting for clustering at community and household levels and associations with key demographic variables assessed through multivariable random-effects logistic regression.

**Results:** 1,318 participants from 477 households were surveyed. The prevalence of scabies prevalence was 9.3% (95% CI: 6.5-13.2%), across 75 (19.7%) households; impetigo or infected scabies prevalence was 0.8% (95% CI: 0.4-1.9%). The majority (52%) of scabies cases were classified as severe. Scabies prevalence was lower in females and higher in the youngest age group; no associations were found with other collected demographic or socio-economic variables. DLQI scores indicated a very or extremely large effect on HRQoL in 29% of adults and 18% of children diagnosed with scabies.

**Conclusions:** Our study indicates a substantial burden of scabies in this peri-Urban population in Liberia. This was associated with significant impact on quality of life, highlighting the need for action to control scabies in this population. Further work is needed to assess the impact of interventions in this context on both the prevalence of scabies and quality of life.

**Plain English summary:** Scabies is an infestation with a microscopic mite which affects many people living in low-resource tropical countries. It causes intense itching, which can lead to complications through bacterial infection and poor quality of life. To help develop global scabies control programmes, we need a better understanding of how common it is across different tropical settings. We conducted a survey to assess the burden of scabies and bacterial skin infection in a random sample of people living in a community in Monrovia, Liberia. Information about participants and their household were collected and their skin was examined; those with skin conditions were asked about its impact on quality of life.

We examined 1,318 participants and found that almost 10% of people had scanies. Scabies was more common in young children, and was more common in male children than female children. We found that there was a large impact on quality of life due mostly to the itching that scabies causes and to people feeling embarrassed or sad because of their skin condition. This scabies survey is one of the first conducted across all age groups in recent years in sub-Saharan Africa and indicates a substantial burden and impact on quality of life. More work is needed to understand how common scabies is in different settings and the impact that different treatment strategies may have.

## Background

Scabies is a parasitic infestation of the skin by the mite *Sarcoptes scabiei* var. *hominis*. The presence of mite products in the stratum corneum drives a host hypersensitivity reaction that results in intense pruritis and rash, which can lead to disrupted sleep, poor concentration and social stigma [1]. Scabies is estimated to affect over 200 million people globally [2] and the highest prevalence is thought to occur in low-resource tropical settings [3]. Scabies can be complicated by secondary bacterial infection with *Streptococcus pyogenes* and *Staphylococcus aureus* [3,4] and the risk of severe conditions including post streptococcal glomerulonephritis, acute rheumatic fever and rheumatic heart disease [3,5]. In view of this burden of disease, scabies was added to the WHO Neglected Tropical Disease portfolio in 2017 [6], with a recommendation that scabies management should be incorporated in the universal health coverage package of care [7].

The development of an effective global control strategy for scabies requires a sound understanding of the disease burden, however robust epidemiological data for the condition is sparse for a number of reasons [8]. Firstly, many settings do not have accurate estimates of scabies prevalence. Routine health data, if collected, often under-estimates the true population burden, arising from reasons including ‘normalisation’ of skin lesions by affected individuals and their under-recognition by clinical staff [9]. A 2015 systematic review of scabies and impetigo prevalence indicated wide variation in prevalence estimates globally; a consistently high burden was seen in many resource-limited settings, particularly in Pacific countries, with the highest prevalence documented in Papua New Guinea at 71% [10]. Prevalence figures from sub-Saharan African countries were lower but few high-quality, population-based data were available. Secondly, until recently there has been a lack of standardised criteria for diagnosis of scabies [10]. The International Alliance for the Control of Scabies (IACS) published consensus criteria for use in both research and public health settings and in particular in low- and middle-income countries [11]. Thirdly, despite associations with stigma and poor mental health [8,12,13], there are limited high-quality data on the impact of scabies and impetigo on health-related quality of life (HRQoL). Improving our understanding of the morbidity associated with scabies is essential in assessing the potential benefit of interventions. Finally, the relationship between scabies and secondary bacterial infection seems to be variable [10], with the latter appearing to be less common in sub-Saharan Africa than in the Pacific, where the majority of studies to date have been conducted [8].

Recently, mass drug administration (MDA) utilising oral ivermectin and 5% permethrin cream has shown promise as an effective control strategy for scabies in areas of high endemicity [14,15]. A greater understanding of the disease burden due to scabies globally is required in order to develop and scale-up control strategies [11]. A recent WHO Informal Consultation on a Framework for Scabies Control identified the need to obtain high-quality population data from regions outside the Pacific, and in particular in peri-urban settings, and to establish mapping methodologies as priorities to assist countries and WHO in developing international guidelines for scabies control. A large outbreak of scabies occurred in Liberia in 2018, including the capital, Monrovia [16]. The Ministry of Health launched measures to control transmission, including treatment of cases and their household contacts. There have been no population-based surveys undertaken since to see if scabies remains a problem in urban and peri-urban settings in the country. We therefore conducted a population-based prevalence survey for scabies, secondary bacterial infection and impetigo in Monrovia, Liberia.

## Materials and methods

### Study setting

The study was conducted in New Kru Town, a peri-urban coastal community in Bushrod Island Monrovia. The area has a population of over 20,000 inhabitants [17] and is composed of 25 distinct communities. New Kru Town was not specifically targeted for scabies intervention during the 2018 scabies outbreak and the underlying burden of infestation in this community is poorly understood.

### Sample size calculation

We assumed an average household size of 4 and an average of 20 households enrolled per community cluster. Based on our predicted average cluster size of 80 and an intraclass correlation coefficients (derived from previous surveys) we assumed a design-effect of 1.8. With a 20% non-participation rate, we needed to enrol 1,200 individuals to have 80% power to detect a prevalence of scabies of 15% with 3% precision. We therefore needed to enrol at least 15 community clusters of 20 households to achieve our sample size.

### Study procedures

We conducted a cross-sectional population-based cluster-randomised household prevalence survey over a 4-week period in February and March 2020. The survey team consisted of three Liberian mid-level health workers (MLHWs) who had been trained to perform skin examinations and diagnose scabies and impetigo and a local guide to accompany them in each community. We have reported on the training methodology and accuracy of the MLHWs compared to a reference standard examination [18].

We randomly selected 15 of the 25 communities in New Kru Town as primary sampling units (Figure 1). An initial community entry meeting was held with the 15 community chairpersons and information about the study was disseminated within each community prior to the survey to enhance local acceptance. In each community a random sample of households was surveyed to achieve a target sample size of 80 participants per community. Households were selected using a random walk method. The centre point of the community was located by a local guide and a random trajectory was determined using a computer-generated random number between 0 and 359. A start point was determined by counting the number of buildings along this trajectory to the community’s edge and using computer-generated random number selection to determine a building number as the start point. From there a random trajectory was chosen and all buildings within 5m to the left were included. When the edge of the community was reached, a further random trajectory was determined to continue the survey until the required sample size was reached. Each building along the trajectory was recorded and every household within an inhabited building was surveyed. If a household was absent, a revisit was arranged for the same day if information on their return could be obtained; when this was not possible, the house was logged as empty and excluded. For the purpose of the survey, a household was defined as a group of people living together who eat from the same pot; all residents in selected households were invited for inclusion.

**Figure 1:**
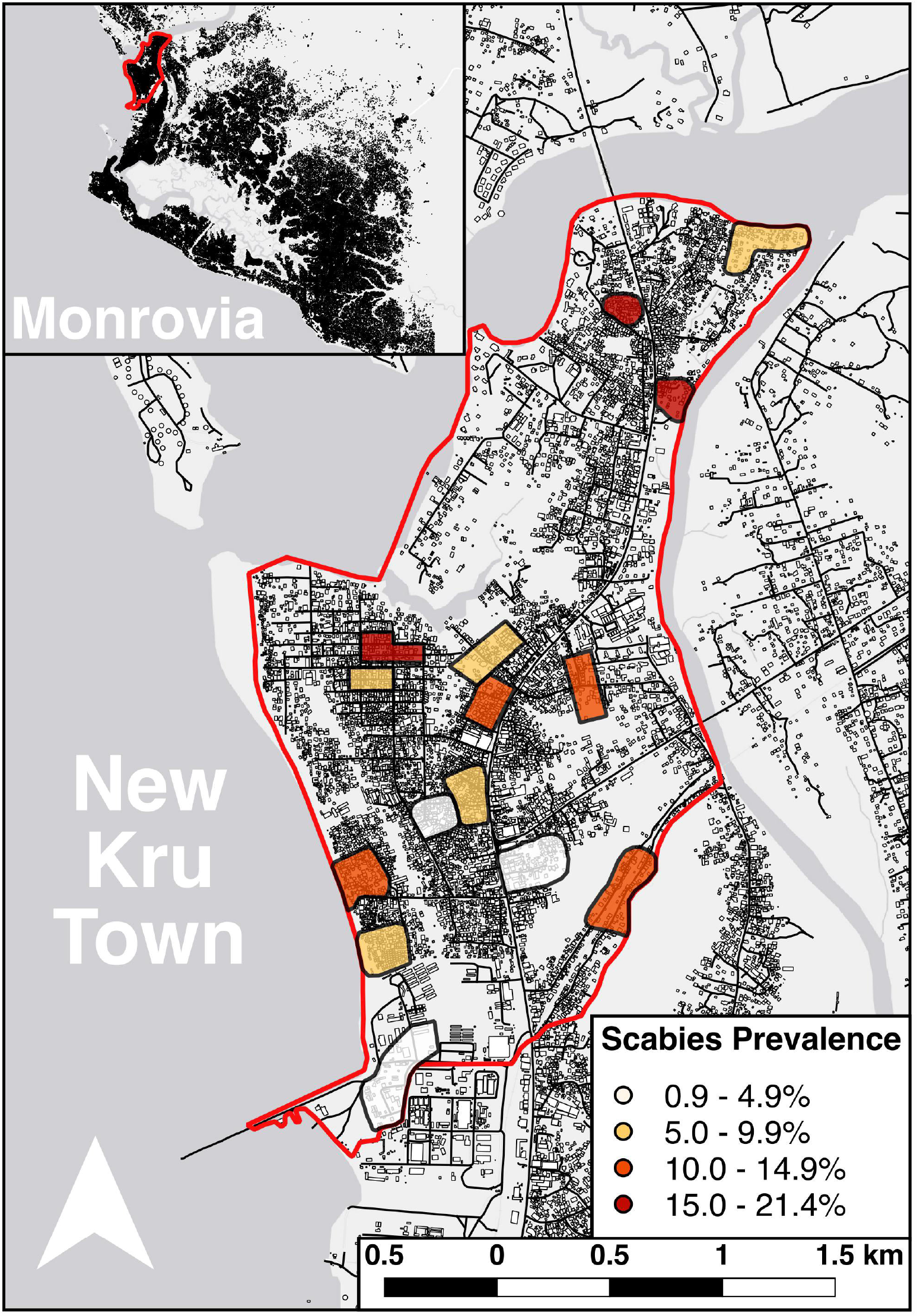
New Kru Town. Underlying map data is derived from OpenStreetMap. The cluster extents are derived from sampling points within the survey.

For each included household we collected data including observed household materials and reported facilities, household size, and demographic details of the household head, including age, gender, level of education and occupation. For each individual, we obtained equivalent demographic details alongside a clinical history and conducted a standardised clinical examination for the detection of scabies, secondary bacterial infection and impetigo. If household members were absent, we arranged a revisit for the same day where practicable; where this was not possible, we recorded their age and gender and logged them as absent. Data were collected electronically on Android devices (Samsung Galaxy Tab A) using OpenDataKit (ODK, Seattle, USA, 2010).

We utilised the IACS consensus criteria for the diagnosis of “clinical” or “suspected” scabies. Scabies was diagnosed if examiners identified burrows (category B1) or found an individual with typical scabies lesions in a typical distribution and one (category C1) or two (category B3) history features [11]. On the basis of our findings from a study of the validity of the IACS criteria and those of a previous study we limited examination to the face, upper limb and axilla and exposed areas of the lower limb for individuals aged five and over, as this has been shown to have a sensitivity of more than 90% for detecting the presence of scabies lesions compared to a full-body examination [18,19], however if an individual reported a lesion elsewhere that they wished to have examined, this was also assessed. A wider distribution of lesions is typically found in infants and therefore children aged less than five years old had a full skin examination performed [11]. All sensitive examinations were performed in private and in the presence of a responsible adult for examinations in children. Impetigo was defined on the basis of erythematous lesions with pus or crust. We classified the extent and severity of scabies and impetigo based on the number of lesions as previously described [19,20].

We assessed HRQoL using a skin disease specific tool in individuals diagnosed with scabies, secondary bacterial infection or impetigo. For individuals aged 16 years and older we utilised the dermatology life quality index (DLQI)[21], for individuals aged seven to 15 years the Children’s DLQI (CDLQI)[22], and for participants under seven a family DLQI (FDLQI)[23] was completed by a parent or responsible adult. We only conducted one FDLQI for each affected household with a child less than seven years old. Individuals who were diagnosed with scabies were offered treatment with oral ivermectin or benzyl-benzoate lotion.

### Statistical analysis

Data were analysed in STATA 16 (StataCorp, Texas, USA, 2019) a R 4.0 (The R Foundation for Statistical Computing). For the purpose of analysis occupation was classified as unemployed, student, market trader, business (other) and other; highest level of education completed was classified as no formal education, primary school education, secondary school incomplete or complete and college/university complete; latrine type was classified as no latrine (open defecation), open pit latrine or pour-flush toilet. Prevalence estimates for scabies, secondary bacterial infection and impetigo were calculated accounting for clustering at the community and household level. Adjusted odds ratios (aOR) were calculated for key demographic variables using a multivariable random-effects logistic regression model adjusted for age and gender and accounting for clustering as above. The relationship between scabies and impetigo and HRQoL was assessed by calculating the DLQI, the CDLQI for children above 7 and the FDLQI for children for below 7.

### Ethics statement

Written informed consent was obtained from participants aged 18 years and older and from the parents or responsible adults for children. Verbal assent was obtained from children who were able to provide it. The study was approved by The London School of Hygiene and Tropical Medicine, UK (Ref 17796) and the University of Liberia – Pacific Institute for Research and Evaluation Institutional Review Board, Liberia (Ref 20-01-195).

## Results

1,318 participants from 477 households, across 15 communities were enrolled (Figure 2). The median age of participants was 21 years (IQR 9-34) and 61% were female. The mean number of people was surveyed in each community was 88 (range 78-106) (Table 1).

**Table 1:**
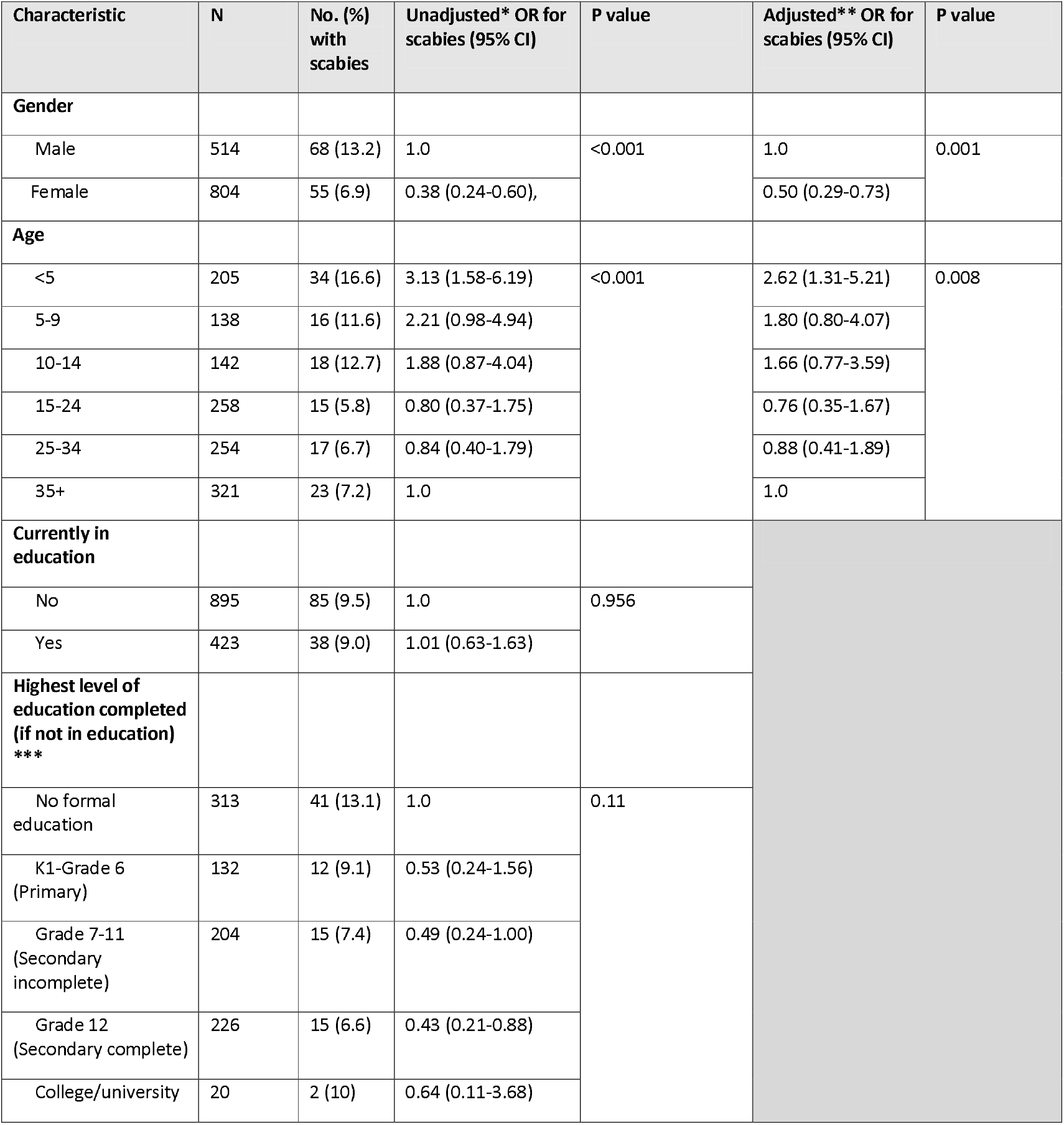
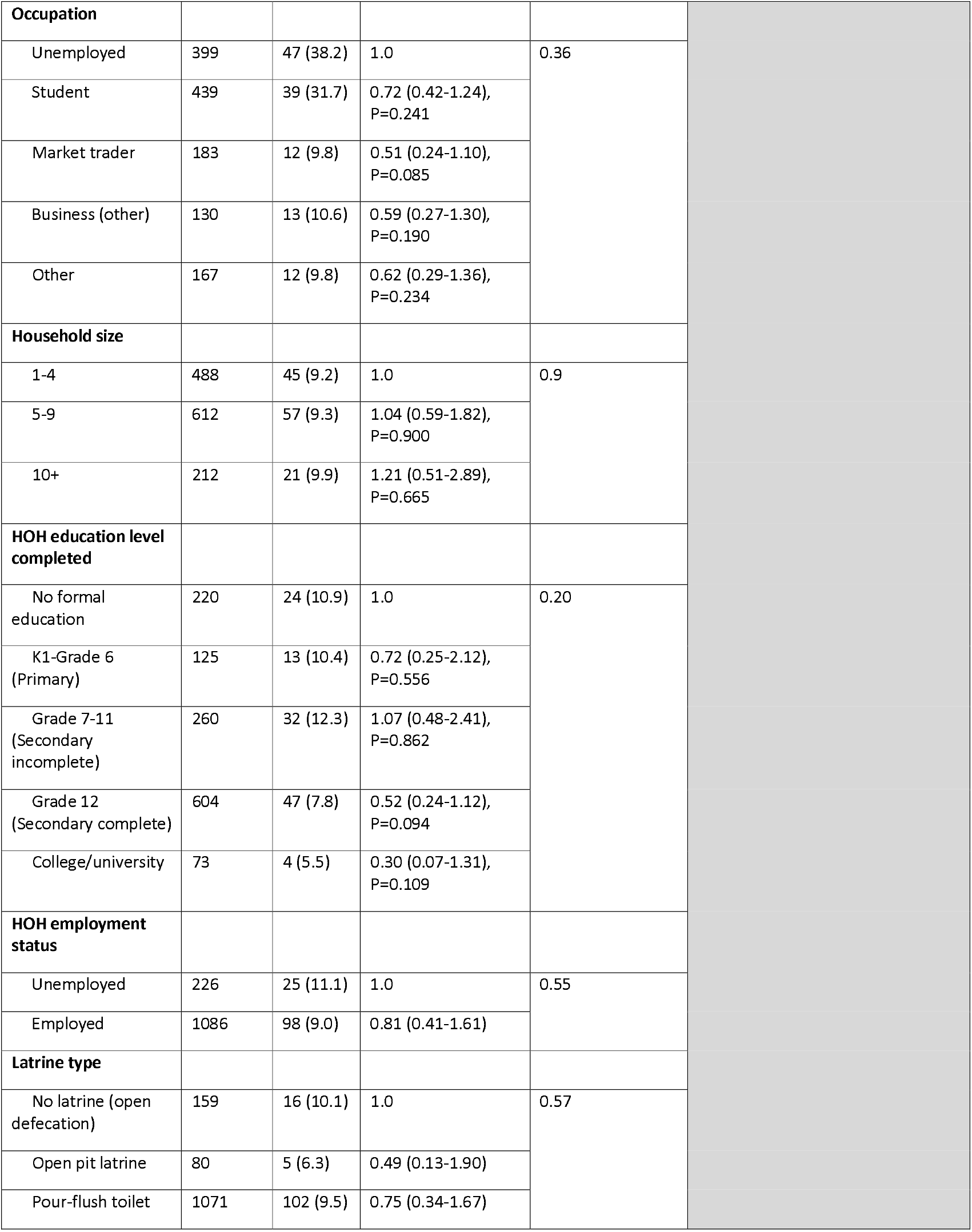
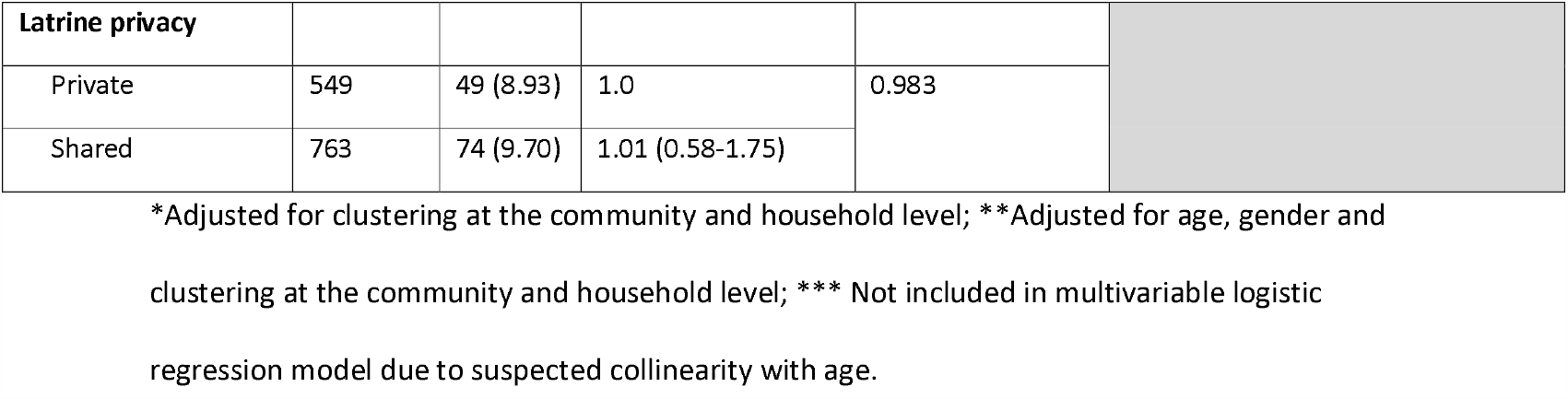
Scabies prevalence by key demographic and household characteristics.

| Characteristic | N | No. (%)<br>with<br>scabies | Unadjusted* OR for<br>scabies (95% CI) | P value | Adjusted** OR for<br>scabies (95% CI) | P value |
| --- | --- | --- | --- | --- | --- | --- |
| Gender |  |  |  |  |  |  |
| Male | 514 | 68 (13.2) | 1.0 | <0.001 | 1.0 | 0.001 |
| Female | 804 | 55 (6.9) | 0.38 (0.24-0.60), |  | 0.50 (0.29-0.73) |  |
| Age |  |  |  |  |  |  |
| <5 | 205 | 34 (16.6) | 3.13 (1.58-6.19) | <0.001 | 2.62 (1.31-5.21) | 0.008 |
| 5-9 | 138 | 16 (11.6) | 2.21 (0.98-4.94) |  | 1.80 (0.80-4.07) |  |
| 10-14 | 142 | 18 (12.7) | 1.88 (0.87-4.04) |  | 1.66 (0.77-3.59) |  |
| 15-24 | 258 | 15 (5.8) | 0.80 (0.37-1.75) |  | 0.76 (0.35-1.67) |  |
| 25-34 | 254 | 17 (6.7) | 0.84 (0.40-1.79) |  | 0.88 (0.41-1.89) |  |
| 35+ | 321 | 23 (7.2) | 1.0 |  | 1.0 |  |
| Currently in<br>education |  |  |  |  |  |  |
| No | 895 | 85 (9.5) | 1.0 | 0.956 |  |  |
| Yes | 423 | 38 (9.0) | 1.01 (0.63-1.63) |  |  |  |
| Highest level of<br>education completed<br>(if not in education)<br>*** |  |  |  |  |  |  |
| No formal<br>education | 313 | 41 (13.1) | 1.0 | 0.11 |  |  |
| K1-Grade 6<br>(Primary) | 132 | 12 (9.1) | 0.53 (0.24-1.56) |  |  |  |
| Grade 7-11<br>(Secondary<br>incomplete) | 204 | 15 (7.4) | 0.49 (0.24-1.00) |  |  |  |
| Grade 12<br>(Secondary complete) | 226 | 15 (6.6) | 0.43 (0.21-0.88) |  |  |  |
| College/university | 20 | 2 (10) | 0.64 (0.11-3.68) |  |  |  |

|  |  |  |  |  |
| --- | --- | --- | --- | --- |
| **Occupation** |  |  |  |  |
| Unemployed | 399 | 47 (38.2) | 1.0 | 0.36 |
| Student | 439 | 39 (31.7) | 0.72 (0.42-1.24),<br>P=0.241 |  |
| Market trader | 183 | 12 (9.8) | 0.51 (0.24-1.10),<br>P=0.085 |  |
| Business (other) | 130 | 13 (10.6) | 0.59 (0.27-1.30),<br>P=0.190 |  |
| Other | 167 | 12 (9.8) | 0.62 (0.29-1.36),<br>P=0.234 |  |
| **Household size** |  |  |  |  |
| 1-4 | 488 | 45 (9.2) | 1.0 | 0.9 |
| 5-9 | 612 | 57 (9.3) | 1.04 (0.59-1.82),<br>P=0.900 |  |
| 10+ | 212 | 21 (9.9) | 1.21 (0.51-2.89),<br>P=0.665 |  |
| **HOH education level completed** |  |  |  |  |
| No formal education | 220 | 24 (10.9) | 1.0 | 0.20 |
| K1-Grade 6 (Primary) | 125 | 13 (10.4) | 0.72 (0.25-2.12),<br>P=0.556 |  |
| Grade 7-11 (Secondary incomplete) | 260 | 32 (12.3) | 1.07 (0.48-2.41),<br>P=0.862 |  |
| Grade 12 (Secondary complete) | 604 | 47 (7.8) | 0.52 (0.24-1.12),<br>P=0.094 |  |
| College/university | 73 | 4 (5.5) | 0.30 (0.07-1.31),<br>P=0.109 |  |
| **HOH employment status** |  |  |  |  |
| Unemployed | 226 | 25 (11.1) | 1.0 | 0.55 |
| Employed | 1086 | 98 (9.0) | 0.81 (0.41-1.61) |  |
| **Latrine type** |  |  |  |  |
| No latrine (open defecation) | 159 | 16 (10.1) | 1.0 | 0.57 |
| Open pit latrine | 80 | 5 (6.3) | 0.49 (0.13-1.90) |  |
| Pour-flush toilet | 1071 | 102 (9.5) | 0.75 (0.34-1.67) |  |

| Latrine privacy |  |  |  |  |
| --- | --- | --- | --- | --- |
| Private | 549 | 49 (8.93) | 1.0 | 0.983 |
| Shared | 763 | 74 (9.70) | 1.01 (0.58-1.75) |  |

\*Adjusted for clustering at the community and household level; \*\*Adjusted for age, gender and clustering at the community and household level; \*\*\* Not included in multivariable logistic regression model due to suspected collinearity with age.

**Table 2:** Prevalence of scabies and impetigo and scabies diagnostic characteristics.

| **Prevalence of disease in study population** | **N (%; 95%CI)** |
| --- | --- |
| **Scabies** | **123 (9.3, 6.5-13.2)** |
| **Secondary Bacterial Infections** | **11 (0.7, 0.3-1.3)** |
| **Impetigo** | **2 (0.2, 0.03-0.6)** |
| **Scabies severity** |  |
| Mild scabies (1-10 lesions) | 21 (17.1) |
| Moderate scabies (11-49 lesions) | 38 (30.9) |
| Severe scabies (50+ lesions) | 64 (52) |
| **IACS criteria** |  |
| B1 | 1 (0.8) |
| B3 | 94 (76.4) |
| C1 | 28 (22.8) |

**Figure 2:**
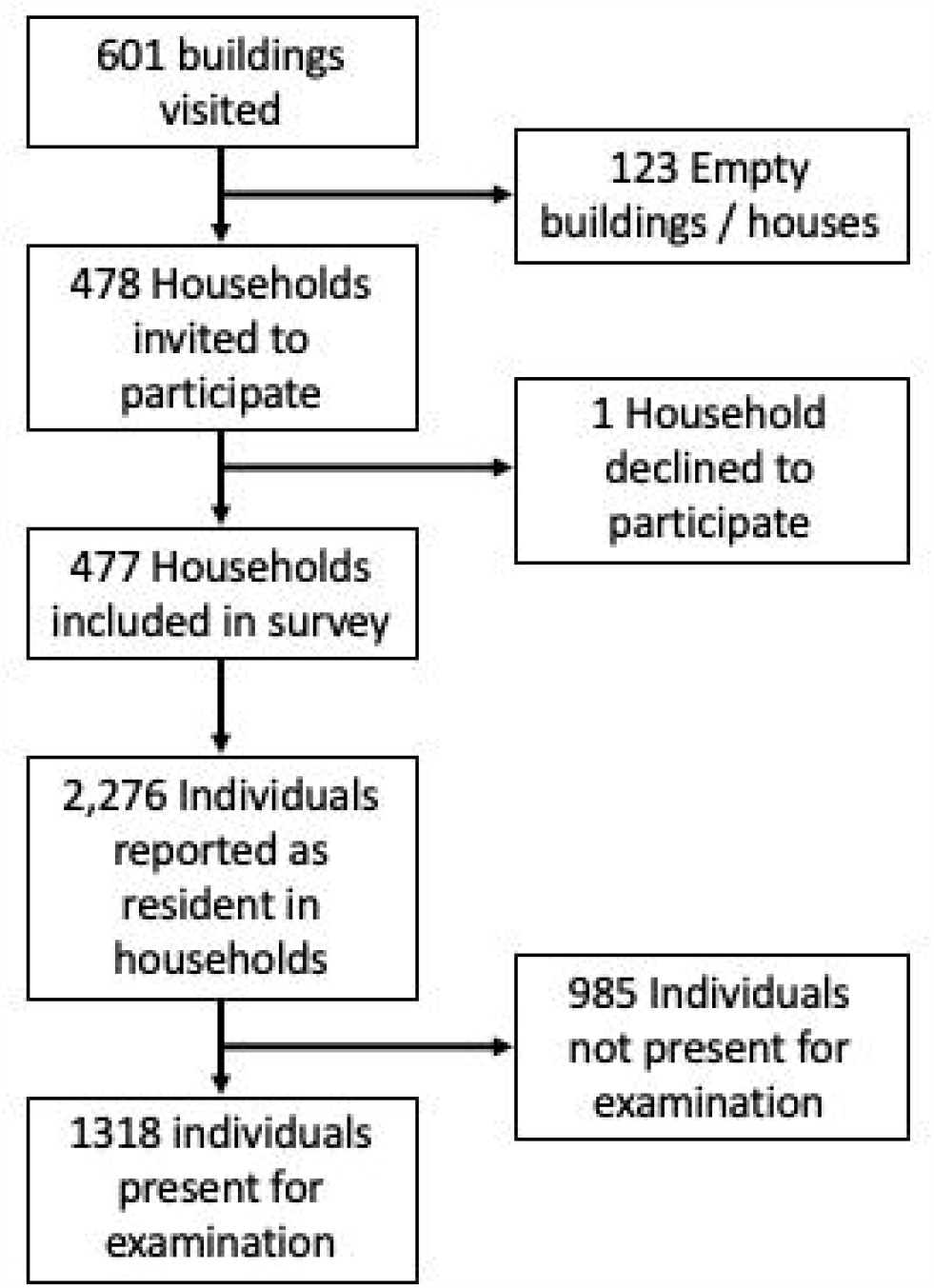
Study enrolment flowchart.

The prevalence of scabies across the 15 communities was 9.3% (N=123, 95% CI: 6.5-13.2%), with a range by community of 0.9-21.4% and the overall prevalence of impetigo or infected scabies was 0.8% (N=11, 95% CI: 0.4-1.9%). Cases of scabies were found in 75 of 380 households (19.7%). Of 11 patients diagnosed with impetigo, nine also had a diagnosis of scabies (Table 1). A large majority of scabies cases (76.4%) were classified as clinical scabies (IACS category B3). Based on the number of lesions, over half of scabies cases were classified as severe (52%). Overall, the prevalence of scabies was lower in females compared to males (6.9% vs 13.2%, aOR 0.50, 95% CI 0.29-0.73), this difference was due to higher prevalence in male children than female children. The prevalence was also higher in the youngest age group compared to the oldest, with 16.6% of those under five diagnosed with scabies compared to 7.2% of those aged 35 and older (aOR 2.62, 95% CI 1.31-5.21). We did not find that any other collected demographic or socio-economic variables were associated with scabies (Table 1) but there was evidence of clustering at the community level (ICC 0.1 95% CI 0.04 – 0.26, p <0.001).

Across the overall study population, 484 (36.7%) people reported having itch. The sensitivity of self-reported itch for an eventual diagnosis of scabies was 97.6% with a specificity of 69.5%. A similar picture was seen at household level, with a sensitivity of any individual in a house reporting itch for a case of scabies in the house of 98.5% and a specificity of 56.2%. At the cluster level there was a moderate correlation between the prevalence of itch and the prevalence of scabies (r = 0.6) (Table 3).

**Table 3:** Association between scabies diagnosis and self-reported history of itch.

| **Participant-reported itch history (N=1318)** | **N (%) survey population** | **N (%) scabies cases** | **Sensitivity for scabies diagnosis** | **Specificity for scabies diagnosis** | **PPV** | **NPV** |
| --- | --- | --- | --- | --- | --- | --- |
| Personal history of itch | 484 (36.7) | 120 (97.6) | 97.6%<br>(92.5 – 99.4%) | 69.5%<br>(66.8 – 72.1%) | 24.8%<br>(21.1 – 28.9%) | 99.6%<br>(98.9 – 99.9%) |
| Household member with itch | 488 (37) | 97 (78.9) | 78.9%<br>(70.4 – 85.5%) | 67.3%<br>(64.5 – 69.9%) | 19.9%<br>(16.5 – 23.8%) | 96.9%<br>(95.4 – 97.9%) |
| Other contact with itch | 104 (7.9) | 12 (9.8) | 9.8%<br>(5.4 – 16.8%) | 92.3%<br>(90.6-93.7%) | 11.5%<br>(6.4-19.7%) | 90.9%<br>(89.1 – 92.4%) |
| **Household-level itch history (N=477)** | **N (%) households** | **N (%) households with a scabies case** | **Sensitivity for scabies diagnosis in household** | **Specificity for scabies diagnosis in household** | **PPV** | **NPV** |
| Household member reporting itch | 257 | 86 | 98.5%<br>(92.9 – 99.9%) | 56.2%<br>(51.1 – 61.1%) | 33.5%<br>(28.2 – 40.0%) | 99.5%<br>(97.1 – 99.9%) |

A HRQoL measure was completed with all participants (or their responsible adult where appropriate) diagnosed with scabies and/or impetigo. The median score among adults completing the DLQI was 8 (IQR 4-11), reflecting a moderate effect on HRQoL, with 29% of adults achieving scores which indicated a very or extremely large effect on quality of life. Among children aged 7 years and older completing the CDLQI, the median score was 6 (IQR 4-10), reflecting a small effect on quality of life, with 18% achieving scores which indicated a very or extremely large effect. The median score of the FDLQI was 7 out of a possible score of 30 (IQR 6-9) consistent with a moderate impact on quality of life. Higher DLQI/CDQLI scores were driven predominantly by higher scores on questions related to itch and feelings of embarrassment or low mood related to their skin. There was evidence that the severity of disease, as measured by lesion counts, was associated with worse quality of life outcomes (Table 4).

**Table 4:** DLQI scores of scabies cases and stratification by scabies severity.

| **DLQI score (N=58)** | **N (%) of cases** | **N (%) mild scabies** | **N (%) moderate scabies** | **N (%) severe scabies** |
| --- | --- | --- | --- | --- |
| 0-1 (no effect on patient's life) | 4 (6.9) | 3 (75) | 0 (0) | 1 (25) |
| 2-5 (small effect) | 16 (27.6) | 5 (31) | 7 (44) | 4 (25) |
| 6-10 (moderate effect) | 21 (36.2) | 4 (19) | 7 (33) | 10 (48) |
| 11-20 (very large effect) | 15 (25.9) | 1 (7) | 4 (27) | 10 (67) |
| 21-30 (extremely large effect) | 2 (3.5) | 0 (0) | 0 (0) | 2 (100) |
| **CDLQI score (N=28)** |  |  |  |  |
| 0-1 (no effect on child's life) | 2 (7.1) | 0 (0) | 2 (100) | 0 (0) |
| 2-6 (small effect) | 13 (46.4) | 3 (23) | 7 (54) | 3 (23) |
| 7-12 (moderate effect) | 8 (28.6) | 1 (13) | 1 (13) | 6 (75) |
| 13-18 (very large effect) | 4 (14.3) | 0 (0) | 1 (25) | 3 (75) |
| 19-30 (extremely large effect) | 1 (3.6) | 0 (0) | 0 (0) | 1 (100) |
| **FDLQI score (N=38), median (IQR)** | 7/30 (6-9)) |  |  |  |

## Discussion

This community-based study conducted in peri-urban Monrovia is one of the first scabies prevalence surveys to have been conducted across all age groups in recent years in sub-Saharan Africa and is the first to utilise the 2020 IACS criteria for the diagnosis of scabies. We found an overall prevalence of 9.3% but with a prevalence above 10% amongst children and that this was associated with significant impacts on quality of life, predominantly driven by pruritus and impacts on mental health.

The majority of scabies burden available for sub Saharan Africa have been collected in rural settings. The overall prevalence found in our survey is higher than the 6% found in a rural survey in Tanzania [24] and the 2.8% found in a rural survey in Cameroon [25]; this may in part reflect the increased risk of infection that is thought to arise from overcrowded conditions and which may be more representative of our peri-urban study population [26]. The prevalence of scabies in our survey was found to be higher in younger age groups, which is consistent with other studies [10,24]. The highest burden was found in children under 5 years of age (16.6%), which is at the higher end of prevalence figures seen among this age group in other West African studies [27–31]. However, whilst the highest prevalence was seen among children, the results of our survey indicate that scabies carries a substantial burden across all age groups. We found very strong evidence that the risk of scabies was twice as high in male children compared to females, which has also been reported in some other studies [20], but did not find any evidence of statistical associations between other demographic or socioeconomic characteristics and scabies. It is possible, however, that our study population was too homogeneous to detect an association with these variables.

We did not find a high burden of secondary bacterial infection among the study population, suggesting that this may not be a substantial risk arising from scabies in this setting. We conducted out study in the dry season, and in other West African settings impetigo has been shown to increase markedly in the wet season and we may therefore have underestimated the true burden of impetigo in this setting [27,28]. Regardless, the impact of scabies itself appears to be significant. Over half of all scabies cases diagnosed during the survey (52%) were categorised as extensive, with individuals each having 50 or more lesions. As previously reported, the sensitivity of MLHWs in diagnosing scabies is greater in more extensive disease [18,32] and it is possible that some cases of less extensive disease were not captured, however the number of cases of extensive disease indicates a substantial unmet need in scabies management. Our study further indicates that scabies has a large effect on quality of life. From our validation study, extent of disease, as measured by lesion counts, correlated with the degree of itchiness experienced by the patient [18]. Pruritic skin diseases are known to have a negative impact on quality of life and our data suggests that that pruritus associated with scabies is one of the major drivers of the impact of infestation on HRQoL scores. Though the numbers were too small for formal analysis, those scoring in the highest three categories on the DLQI and CDLQI were most commonly those with extensive disease as measured by lesion counts. It is possible therefore that measures to reduce the burden of scabies would bring substantial benefit to individuals.

The burden of scabies found in this peri-urban community is substantial and highlights that further consideration should be given to population-level management. The 2019 WHO Informal Consultation for a scabies control programme posited that communities with prevalence rates ≥10% should be considered for MDA programmes [33], however there is currently no formal guidance and further research is needed to inform the development of scabies control strategies. Whilst itchiness was highly sensitive for the diagnosis of scabies it had extremely poor specificity at both the individual and the household level suggesting that it is unlikely to a useful marker for guiding decision. Further work is needed to assess whether MDA would have a sustained impact in reducing the prevalence of scabies in sub-Saharan African settings, as has been the case in studies in Fiji [14,34] and to understand what would be the most appropriate treatment to use.

### Limitations

Our study has some limitations. Firstly, not all individuals were present at the time of examination and this may have affected our overall prevalence estimate especially if presence or absence was associated with scabies. Secondly, the sensitivity of MLHWs is lower than an expert examiner which is likely to have resulted in under-estimation of the true burden of disease. The MLHWs who undertook the examinations in this survey underwent training and validation as part of our overall study in Liberia and were found to have similar performance to staff who have undertaken surveys in many other studies, so we do not believe this is a major limitation [18]. Thirdly, our study was conducted in a single community within Monrovia and may not be generalisable to other peri-urban settings. Finally, the DLQI, CDLQI and FDLQI have not been validated for use in Liberia. A consensus was achieved among the MLHW for minor adjustments to the phrasing of questions that would make them appropriate for use in this setting and it was agreed that the content of the questions was still applicable. Further work to formally validate these tools in similar settings would provide increased confidence of findings in future studies, enhancing understanding of the full burden of common skin conditions in these settings.

## Conclusions

Our study highlights that scabies carries a substantial burden across all age groups in this peri-urban community in Monrovia, including a considerable impact on HRQoL for those with the disease. The robust methodology used provides high quality community-level data which makes an important contribution to our understanding of the burden of scabies and impetigo in low resource settings. Further work is needed to increase our understanding of scabies epidemiology across different settings and regions in sub-Saharan Africa. The impact of disease management strategies, including MDA, on scabies prevalence in these settings is essential to inform the development of effective population-level scabies control programmes.

## Data Availability

Data are available in the supplementary data file

## Acknowledgements

The authors would like to thank the participants and members of the New Kru Town community in Monrovia.

---

**Supplementary Dataset 1: Study Dataset**

